# Prevalence and Detection of Obstructive Sleep Apnea Early after Stroke

**DOI:** 10.1101/2024.06.16.24309011

**Authors:** Karen J. Klingman, Sandra A. Billinger, Amanda Britton-Carpenter, Bria Bartsch, Pamela W. Duncan, George D. Fulk

## Abstract

**Background:** Obstructive sleep apnea (OSA) negatively impacts post-stroke recovery. This study’s purpose: examine the prevalence of undiagnosed OSA and describe a simple tool to identify those at-risk for OSA in the early phase of stroke recovery.

**Methods:** This was a cross-sectional descriptive study of people ∼15 days post-stroke. Adults with stroke diagnosis admitted to inpatient rehabilitation over a 3-year period were included if they were alert/arousable, able to consent/assent to participation, and excluded if they had a pre-existing OSA diagnosis, other neurologic health conditions, recent craniectomy, global aphasia, inability to ambulate 150 feet independently pre-stroke, pregnant, or inability to understand English. OSA was deemed present if oxygen desaturation index (ODI) of >=15 resulted from overnight oximetry measures. Prevalence of OSA was determined accordingly. Four participant characteristics comprised the “BASH” tool (body mass index >=35, age>=50, sex=male, hypertension=yes). A receiver operator characteristics (ROC) curve analysis was performed with BASH as test variable and OSA presence as state variable.

**Results:** Participants (n=123) were 50.4% male, averaged 64.12 years old (sd 14.08), and self-identified race as 75.6% White, 20.3% Black/African American, 2.4%>1 race, and 1.6% other; 22% had OSA. ROC analysis indicated BASH score >=3 predicts presence of OSA (sensitivity=0.778, specificity=0.656, area under the curve =0.746, p<0.001).

**Conclusions:** Prevalence of undiagnosed OSA in the early stroke recovery phase is high. With detection of OSA post-stroke, it may be possible to offset untreated OSA’s deleterious impact on post-stroke recovery of function. The BASH tool is an effective OSA screener for this application.

---

Obstructive sleep apnea (OSA) is both a risk factor and consequence of stroke ^1–3^, and negatively impacts recovery and morbidity in people following stroke ^4,5^. The reported prevalence of OSA among people with stroke is high, ranging from 40-72% ^6–9^. Poor sleep, such as that arising from untreated OSA, interferes with memory consolidation, which is a critical factor for the process of motor learning during post-stroke rehabilitation ^10–12^. Further, poor sleep has been linked to less favorable post-stroke outcomes such as worse recovery of function, more disability, longer hospital stays, and poor quality of life. ^13–17^.

Further, there is a growing body of evidence that adverse effects of OSA-related poor sleep on functional recovery can be mitigated by providing treatment for OSA during the acute recovery phase of stroke, and lead to more favorable post-stroke outcomes than if OSA were left untreated ^6,18,19^. It is important, therefore, to understand the relationships between sleep and stroke recovery, and sleep experts recommend screening people with stroke for OSA ^20,21^.

Several OSA screening tools for people with stroke have demonstrated promising psychometric properties in this population ^8,22^. A number of them are based on the STOP-BANG questionnaire^23^ which assigns one point for each of the following OSA risk factors: snoring (S), feeling tired during the day (T), having been observed to stop breathing while asleep (O), high blood pressure (P) (i.e., hypertension diagnosis), high body mass index (B) (BMI>35), older age (A) (>50 years old), high neck circumference (N) (>40 cm); and male gender (G); these are the STOP-BAG (which does not account for neck circumference), and STOP-BAG-O (which adds a factor based on measured overnight oxygen desaturation to the STOP-BAG). Other tools studied included the 4 variable (“4V”)^24^ questionnaire (comprising four of the STOP-BANG items - sex, BMI, blood pressure and snoring), the Berlin Questionnaire (“BQ”) ^25^ (11 items self-report items about sleep and its daytime consequences), and the sleep obstructive apnea score (“SOS”) optimized for stroke patients ^26^ (19 self-report items about sleep and its daytime consequences). Each of these tools, except the SOS, showed promising psychometric properties, ^8^ ^22^ ^25^.

The available OSA screening tools are simple, requiring minimal physical measurements and/or patient responses to questions. People with stroke, however, are rarely screened for OSA symptoms such as snoring and daytime sleepiness in clinical and rehabilitation settings, or offered testing for OSA in the first few months following stroke; for example, only 6% of people with stroke were offered OSA testing, and only 5% were asked about snoring in the first 90 days post-stroke, according to one study ^27^. OSA screening may be low due to the U. S. Preventive Services Task Force’s lack of support to routinely screen for OSA in the general population ^28^. However, in the stroke population, OSA is a risk factor for poor outcomes and warrants additional attention^20,21^.

The purpose of the present study was to examine the prevalence of undiagnosed OSA in people with stroke at the early phase of recovery (first 15 days following stroke). The study-driving hypothesis was that at least 10% of people with stroke would have previously undetected OSA, defined here as oxygen desaturation index of >=15 events per hour ^29,30^. An additional goal of this study was to utilize results of these observations to describe a simple tool, requiring only information readily available in medical records, that could easily be utilized by providers at inpatient rehabilitation facilities to identify people with stroke who are at risk for OSA.

## Methods

### Study Type and Setting

For this cross-sectional descriptive study we used data covering a three-year timeframe from an on-going observational cohort study designed to determine the prevalence and impact of non-OSA sleep disorders in people with stroke^31^. For the parent study (funded by NIH/NINR - R01NR018979), potential participants are screened for OSA as described below. Only those who screen negative for OSA participate in the rest of the parent study, with data collected at three timepoints: during inpatient rehabilitation (approximately 15 days post-stroke), at 60 days post stroke (at home), and at 90 days post stroke (at home). The parent study comprises multiple inpatient rehabilitation data collection locations in the eastern and midwestern United States. A single institutional review board (sIRB) has approved the parent study. WCG IRB (https://www.wcgclinical.com/solutions/irb-review/) is the sIRB of record; the WCG IRB Protocol Number is 20202548. The present study and this summary adhere to the Strengthening the Reporting of Observational Studies in Epidemiology (STROBE) guidelines^32^.

#### Inclusion and Exclusion Criteria

For the parent study, stroke patients aged 18 and older admitted to inpatient rehabilitation units in the eastern and midwestern United States are approached for participation if they meet these inclusion criteria:

- Diagnosis of stroke.
- Age 18 or older.
- Alert or arousable, i.e., National Institutes of Health Stroke Scale (NIHSS) item 1a score <2)^33^.
- Able to provide informed consent or assent.

People with stroke are not approached for participation if they meet any of these exclusion criteria:

- Pre-existing diagnosis of obstructive sleep apnea (OSA), determined from the medical record.
- Living in a nursing home or assisted living center prior to the stroke.
- Unable to ambulate 150 feet independently prior to the stroke.
- Other neurologic health condition that may impact recovery such as Parkinson Disease, Multiple Sclerosis, Traumatic Brain Injury, Alzheimer’s Disease.
- Women who are pregnant.
- Recent hemicraniectomy or suboccipital craniectomy (i.e., those whose bone has not yet been replaced), or any other recent bone removal procedure for relief of intracranial pressure.
- Planned discharge location >150 miles radius from the recruiting rehabilitation facility.
- Global aphasia as defined by a NIHSS item 9 score of 3^33^.
- Inability to understand English.

#### Categorization of Participants for Present Study

After participants provide informed consent/assent with caregiver consent, the first step of the parent study is to screen participants for OSA according to oxygen desaturation index (ODI) based on overnight oximetry (see “measures” below). Consenting to this first step also includes consenting to the extraction of health-related and demographic information from electronic medical records.

For purposes of the present study, individuals were then classified as follows: (a) excluded for reason other than pre-existing diagnosis of OSA based on review of the medical record, (b) pre-existing diagnosis of OSA based on review of the medical record, (c) declined participation, (d) agreed to participation and screened positive for OSA (ODI >=15), or (e) agreed to participation and screened negative for OSA (ODI <15) (see Figure 1). This cutpoint was selected because ODI is known to correlate with apnea hypopnea index (AHI) and AHI >=15 is considered diagnostic for OSA in absence of knowledge regarding individuals’ self-report of sleep characteristics (such as snoring, choking / gasping, and daytime sleepiness)^34^.

**Figure 1:**
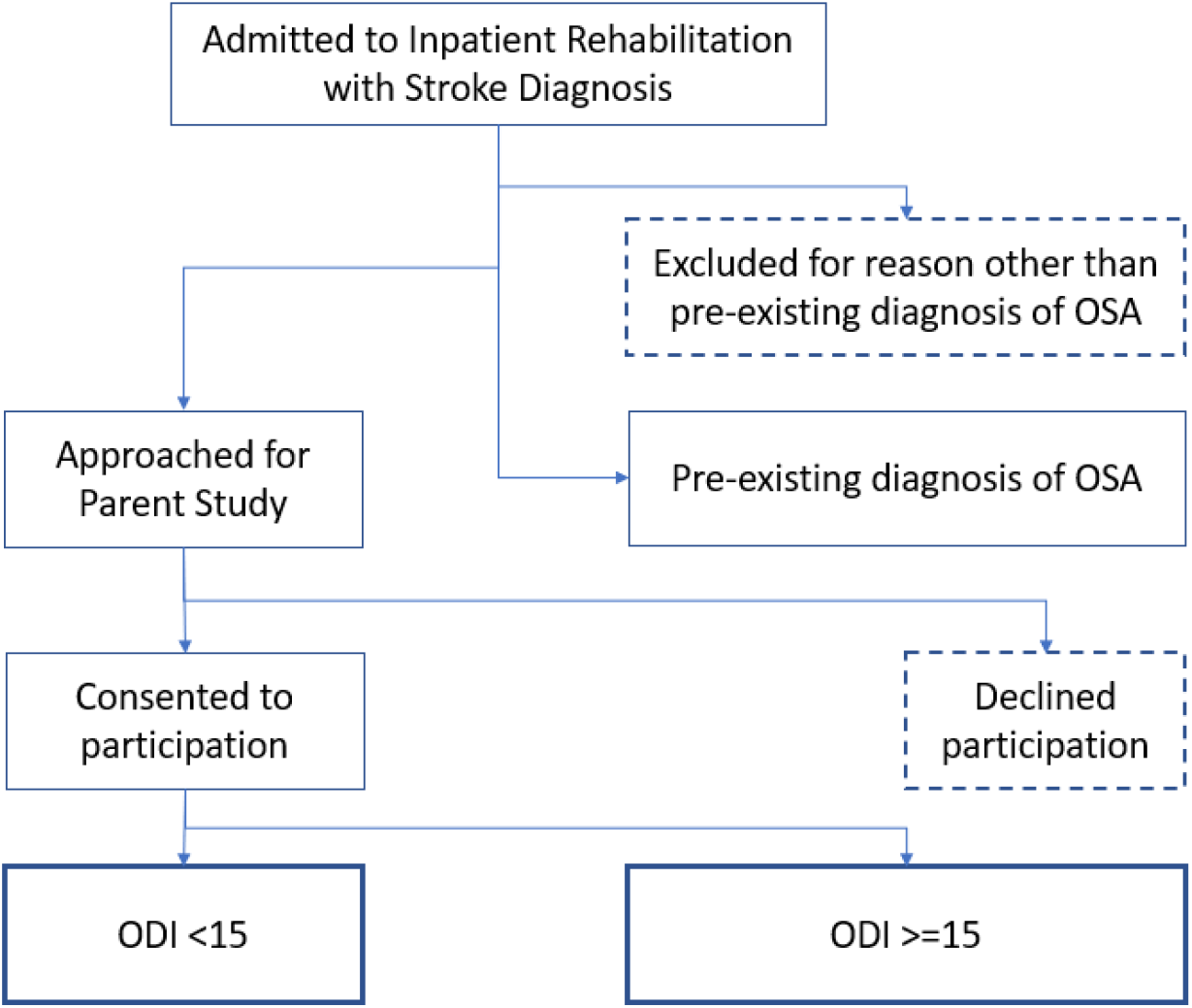
Parent study recruitment flow. *Boxes with dashed lines indicate unknown OSA status*

Individuals who were excluded from the study due to an ODI >=15 were informed of the potential that they might have OSA, and their health care providers were also notified of the oximetry test results.

### Measures

#### Participant Characteristics

Participant characteristics were extracted from medical records. For the present study, demographic information included age and sex. Health-related data included stroke type, stroke location, stroke severity (NIHSS score), height, weight, prior diagnosis of OSA, and prior diagnosis of hypertension.

#### OSA Screening according to Oxygen Desaturation Index (ODI)

Oxygen desaturation index (ODI) has been shown to be an accurate screening metric to detect OSA in patients with stroke ^29,30^. Participants wore a Nonin WristOx2® Model 3150 wrist oximeter (https://www.nonin.com/support/3150-usb/) overnight, and ODI was determined according to the manufacturer’s algorithm. Individuals with ODI >=15 were considered to screen positive for OSA for purposes of the parent study. This cutpoint was selected because ODI is known to correlate with apnea hypopnea index (AHI) and AHI >=15 is considered diagnostic for OSA in absence of knowledge regarding individuals’ self-report of sleep characteristics (such as snoring, choking / gasping, and daytime sleepiness)^34^.

#### “BASH” Screening Measure for OSA

The proposed new screening measure, BASH, includes a subset of elements used to screen for OSA^8,23^. The BASH score ranges from 0-4, and is the sum of its four elements determined as follows:

- B = 1 if **B**ody Mass Index (BMI) >=35, 0 otherwise
- A = 1 if **A**ge >= 50, 0 otherwise
- S = 1 if **S**ex = male, 0 otherwise
- H = 1 if patient has a **H**ypertension diagnosis, 0 otherwise

BMI, age, sex, and hypertension diagnosis were all extracted from the participants’ electronic medical records. The rationale for selecting these elements was to utilize parameters that were readily available in patients’ medical records, without a need for additional physical measurements and query of patients sleep habits. It should also be noted that these four BASH elements are known to be at least partially predictive of OSA, and are a subset of elements utilized by several of the currently available OSA screening tools described earlier (i.e., the STOP-BANG^23^, STOP-BAG-O^8^, STOP-BAG^8^ tools).

### Analyses

#### Participant Characteristics

Demographics, health-related information, BASH scores, and presence of OSA were summarized using descriptive statistics. Numbers of individuals falling into the categories illustrated in Figure 1 were used to determine prevalence of OSA among the study sample.

#### BASH Score vs. OSA Screening Receiver Operating Characteristic (ROC) Analysis

A receiver operating characteristic (ROC) curve analysis was performed to determine practicality of utilizing BASH score to screen for OSA. For purposes of the ROC curve analysis, the test variable was BASH score, and the state variable was based on ODI: state variable =1 (individual screened positive for OSA) if ODI >=15 and state variable = 0 if ODI <15 (individual did not screen positive for OSA). Only individuals with complete sets of data (i.e., no missing data) for the ROC analysis were included. No subgroup analyses were performed.

The practicality of utilizing BASH score as an OSA screening measure was assessed according to area under the curve (AUC) and sensitivity/specificity attained from results of the ROC curve analysis. Values of AUC were interpreted as acceptable (AUC=0.7-0.8), excellent (AUC=0.8-0.9) or outstanding (0.9-1.0)^35^. Sensitivity and specificity were evaluated according to the ways in which false-positives and false-negatives might impact the desirability of implementing BASH as a screening tool for OSA for stroke patients in inpatient rehabilitation facilities.

#### Group Differences in BASH scores

Group differences in BASH scores were determined according to an independent samples t-test between those who did and did not screen positive for OSA. ODI >=15 was considered a positive screen for OSA and ODI <15 was considered a negative screen for OSA. The Cohen’s d resulting from the t-test was utilized to classify the effect size as small (d= 0.2-0.5), moderate (d= 0.5-0.8) or large (d= >=0.8)^36^.

#### Statistical Software

IBM SPSS version 29 was utilized for all statistical analyses.

## Results

### Participant Characteristics

Over a three-year period, n=829 individuals were admitted to participating inpatient rehabilitation facilities with a diagnosis of stroke, and 19% (n=164) had a prior diagnosis of OSA. Of those admitted, 34% (n=286) did not have a pre-existing diagnosis of OSA and met the inclusion/exclusion criteria for the parent study and were approached for participation. Of those approached, 43% (n=123) consented to participate in the parent study, thus yielding n=123 for the analyses to determine the accuracy of the BASH using for ROC curve and group comparison analyses (see Figure 2).

**Figure 2:**
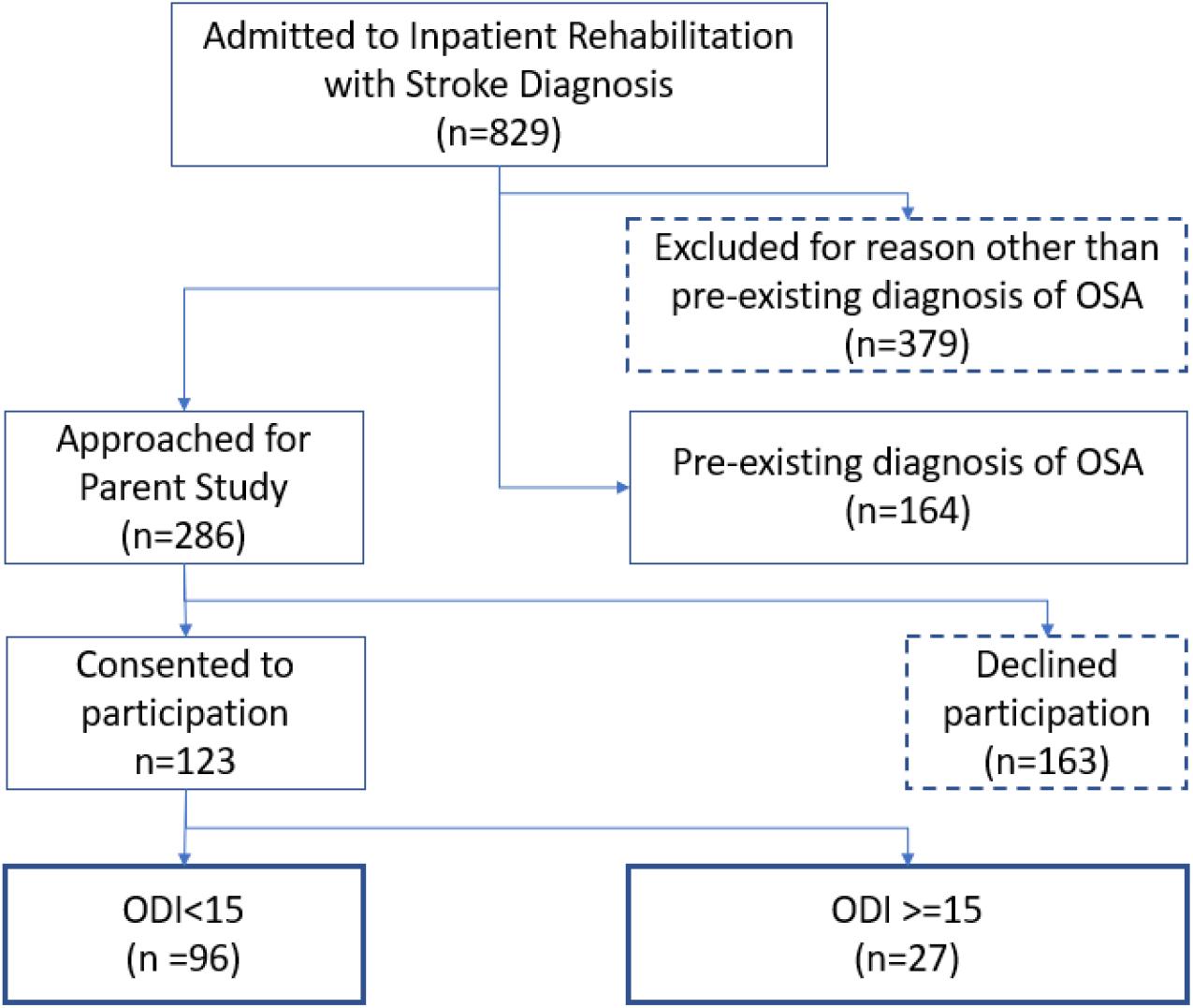
Study participants in each category. The study sample (n=123) was about half (50.4%) male, average age 64.12 (sd 14.08), primarily White (75.6% White, 20.3% Black/African American, 2.4% >1 race, <1% American Indican/Alaskan Native, and <1% unknown), and overweight (average BMI=28.72, sd=6.21); 87% of participants had a diagnosis of hypertension (see Table 1). Stroke severity was moderate (NIHSS average = 5.28, sd=3.41). A majority of the strokes were ischemic (82.1%), with the top three stroke locations being right hemisphere (25.2%), brainstem (20.3%), and right subcortical (19.5%). Other stroke locations observed were were left hemisphere (6.5%), left subcortical (11.4%), cerebellar (9.8%) bilateral (5.7%) and other (1.6%). ODI ranged from 0-60 events/hr (mean ODI=11 sd=12.12), with 22.0% of the participants having an ODI >=15. The time since stroke to ODI measurement averaged 15.28 (sd=7.60) days, and the average BASH score was 2.36, sd=0.75 (see Table 1).

**Table 1:** Participant characteristics.

|  | <u>n</u> | <u>Minimum</u> | <u>Maximum</u> | <u>Mean</u> | <u>s.d.</u> |
| --- | --- | --- | --- | --- | --- |
| Age | 123 | 30.33 | 96.69 | 64.12 | 14.08 |
| BMI | 123 | 17.43 | 52.29 | 28.72 | 6.21 |
| NIHSS total | 105 | 0.00 | 14.00 | 5.28 | 3.41 |
| ODI | 123 | 0.00 | 59.80 | 11.02 | 12.12 |
| Post-stroke days for ODI | 123 | 4.00 | 50.00 | 15.28 | 7.60 |
| BASH score | 123 | 0.00 | 4.00 | 2.36 | 0.75 |
| Sex | <u>n</u> | <u>%</u> |  |  |  |
| M | 62 | 50.4% |  |  |  |
| F | 61 | 49.6% |  |  |  |
| Race |  |  |  |  |  |
| AI/AN<sup>a</sup> | 1 | 0.8% |  |  |  |
| B/AA<sup>b</sup> | 25 | 20.3% |  |  |  |
| White | 93 | 75.6% |  |  |  |
| >1 race | 3 | 2.4% |  |  |  |
| Not reported | 1 | 0.8% |  |  |  |
| Stroke type |  |  |  |  |  |
| Ischemic | 101 | 82.1% |  |  |  |
| Hemorrhagic | 21 | 17.1% |  |  |  |
| Unknown | 1 | 0.8% |  |  |  |
| Stroke location |  |  |  |  |  |
| L hemi | 8 | 6.5% |  |  |  |
| R hemi | 31 | 25.2% |  |  |  |
| L subcort | 14 | 11.4% |  |  |  |
| R subcort | 24 | 19.5% |  |  |  |
| Brainstem | 25 | 20.3% |  |  |  |
| Cerebellar | 12 | 9.8% |  |  |  |
| Bilateral | 7 | 5.7% |  |  |  |
| Other | 2 | 1.6% |  |  |  |
| BASH factors |  |  |  |  |  |
| BMI >= 35 | 18 | 14.6% |  |  |  |
| Age >= 50 | 103 | 83.7% |  |  |  |
| Sex = male | 62 | 50.4% |  |  |  |
| Hypertension | 107 | 87.0% |  |  |  |
| OSA screen (ODI>=15) |  |  |  |  |  |
| No | 96 | 78.0% |  |  |  |
| Yes | 27 | 22.0% |  |  |  |

### BASH Score vs. OSA Screening Receiver Operating Characteristic (ROC) Analysis

The ROC curve analysis of BASH score versus OSA screening status (positive if ODI>=15, negative if <15) yielded an acceptable area under the curve (AUC) of 0.746, p<0.001 (see Figure 3). The point of inflection (maximum Youden index) yielded a BASH cutpoint of >=3 to indicate a positive OSA screen, with sensitivity of 0.778 and specificity of 0.656 (Table 2). At the cutpoint, the likelihood ratios, LR+ and LR-, are 2.26 and 0.336, respectively. These numbers indicate that a person with stroke entering inpatient rehabilitation is 2.26 times more likely to have a positive OSA screen according to BASH score compared to someone without OSA, and 0.336 times as likely to have a negative OSA screen according to BASH score compared to someone without OSA ^37^.

**Figure 3:**
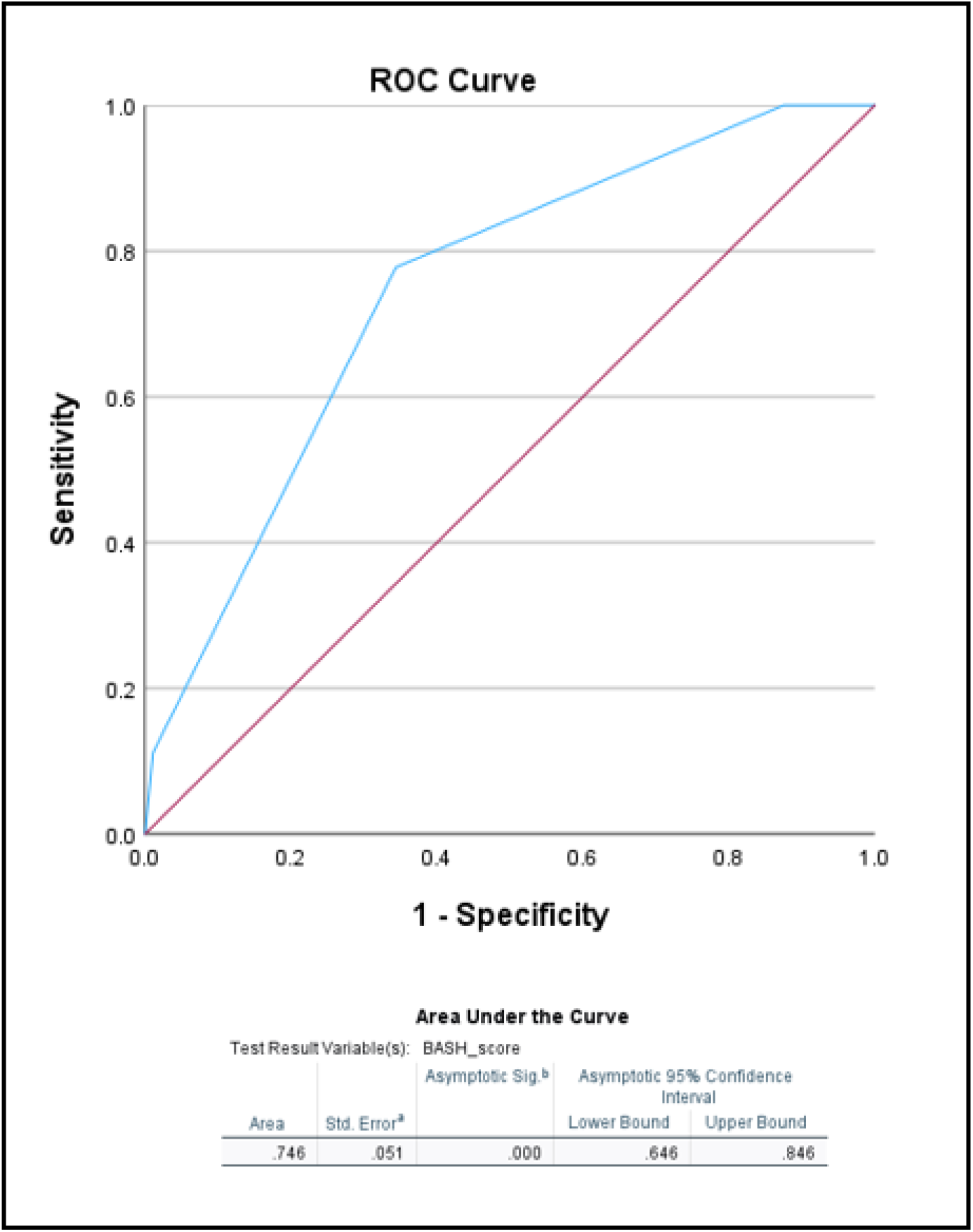
ROC analysis curve.

**Table 2:**
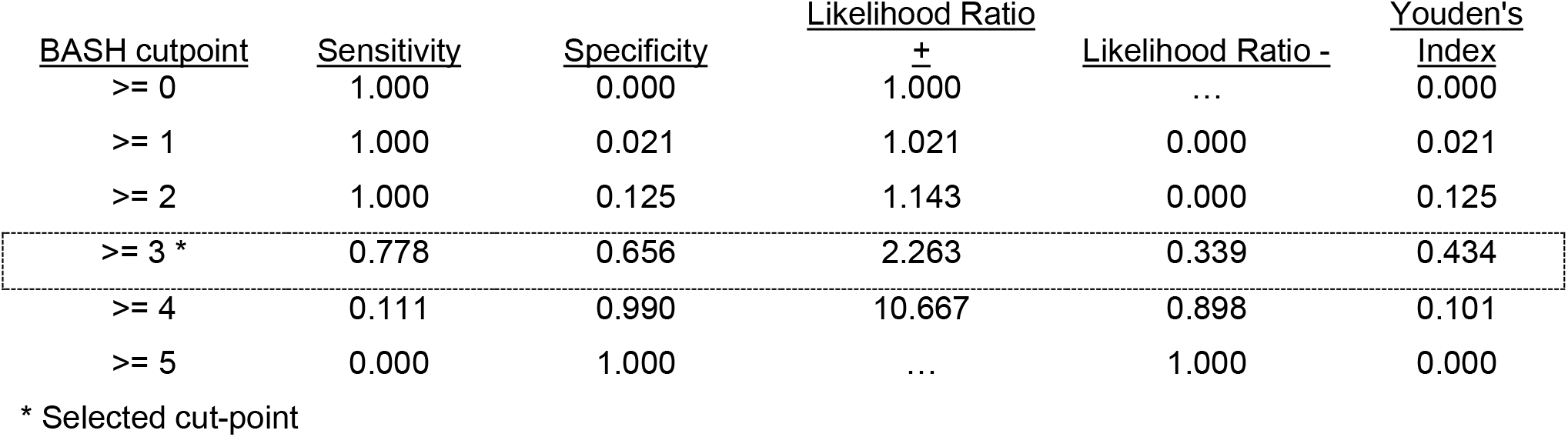
Coordinates of the ROC curve.

### Group Differences in BASH scores

Results of the t-test indicate that there was a significant difference in BASH scores, with large effect size, between participants who screened positive for OSA with an ODI >=15, t(121)= 4.49, p<0.001, Cohen’s d=0.979. The mean BASH score for those who screened positive for OSA (n=27) was 2.89 (sd=0.58) and 2.21 (sd=0.72) for those who screened negative for OSA (n=96).

## Discussion

The BASH tool demonstrated effective properties as an OSA screener for people with stroke during inpatient rehabilitation and should be part of comprehensive post-acute stroke evaluation. In the present study, the high rate of OSA-positive participants with an ODI >=15 (22%) indicates there may missed opportunity to improve the course of stroke recovery unless people with stroke are screened for OSA. Without OSA screening as part of usual care, people with stroke may progress through rehabilitation with untreated OSA, and thus experience poorer recovery outcomes than they could have. The OSA prevalence findings found here are similar to those of Finkel and colleagues^38^ who found that 18.5% of people undergoing surgery had undiagnosed OSA and Alonderis^39^ and colleagues who found that 35% of people with coronary artery disease had undiagnosed OSA. Routine testing for OSA during inpatient rehabilitation is not currently part of the standard of post-stroke care and may add another step to post-stroke inpatient rehabilitation. However, the proposed BASH tool (based on patient characteristics BMI, age, sex, and hypertension diagnosis), provides a quick and easy method for identifying those who should be screened for OSA via overnight oximetry (and potentially referred for additional sleep testing).

Results of this study also confirm a high prevalence of diagnosed OSA among people with stroke, i.e., 20% (n=164 out of N=829, see Figure 2) of all people with stroke admitted to the participating inpatient rehabilitation units had diagnoses of pre-existing OSA in their medical records. Considering the higher population estimates as mentioned above (40-72% ^6–9^), there is concern that OSA is being severely under-detected, even after adjusting for the n=27 positive OSA screens detected during this study. It may be that the individuals who were not included in the analyses for this study disproportionately had OSA. This includes about 65% of the admitted patients with stroke: those who were initially excluded for reasons other than presence of OSA diagnosis in medical record (n=379) or those who declined participation in the parent study (n=155) or dropped prior to overnight oximetry (n=8). It may also be that the sample available for this (and the parent) study was not fully representative of stroke patients due to COVID-19 pandemic-related operational adjustments made by the acute care facilities.

Regardless of the wide range for prevalence of OSA among stroke patients, the low-end estimates indicate that at least one in every five individuals recovering from stroke suffer from OSA. The potential ease of use of the BASH to identify patients with stroke who have OSA suggests it could be used as part of a chart-based algorithm to design post-stroke rehabilitation regimens. Detecting, and therefore explicitly attending to OSA during post-stroke rehabilitation, could lead to improved functional outcomes for the large percentage of patients with stroke with OSA.

The ability of the BASH to distinguish between participants with an ODI less than 15 and greater than or equal to 15 was similar to those of Boulos et al. In people with subacute stroke (approximately 90 days post stroke, they found that the STOP-BAG demonstrated an AUC of 0.685 with a sensitivity of 0.606 and specificity of 0.630 and the STOP-BAG-O demonstrated an AUC of 0.736, sensitivity of 0.864, and specificity of 0.467. Our BASH demonstrated an AUC of 0.746, sensitivity of 0.778 and specificity of 0.656.

### Limitations

There are some limitations to our study. The standard to which we compared the BASH score was ODI determined by overnight nocturnal oximetry, not the gold standard of AHI determined from polysomnography. However, performing polysomnography was cost prohibitive for the parent study. Additionally, as discussed above, we cannot determine if the participants who screened positive for OSA had OSA prior to their stroke or they developed it since their stroke. We have no way of knowing if they followed up for additional testing and whether any additional testing confirmed what the ODI predicted. Further research could also be done earlier after stroke while they are in the acute care hospital. It is possible that an even greater number of people with stroke have undetected OSA the immediate acute (day 1) stage of recovery. Not all people with stroke go on to inpatient rehabilitation.

It should also be noted that, for more than half of the stroke patients admitted to inpatient rehabilitation over the study timeframe (those who were excluded from the parent study), OSA status remained unknown.

### Implications

Given the prevalence and impact of untreated OSA in stroke patients, it is suggested that inpatient rehabilitation facilities include screening for OSA in their patient population, as suggested by sleep experts^20,21^. One approach to OSA screening of people with stroke who are undergoing inpatient rehabilitation could be to implement a process whereby all incoming patients undergo overnight oximetry for determination of ODI. However, our findings indicate that testing all patients may not be necessary. The efficiency of care could be improved by testing only a subset of incoming patients. For example, using a BASH score of >=3 as a trigger to identify patients for who may require additional testing for OSA (e.g., overnight oximetry) could provide a streamlined process to maximize the benefit of post-stroke recovery, while minimizing the risk of not identifying those who might need it. The rates of true positives and false negatives associated with the BASH test could enable identification of most individuals who may have screened positive for OSA according to overnight oximetry, while potentially missing only a handful. There may also be a moderate percentage of individuals denoted as positive for OSA according to BASH who screen negative for OSA according to oximetry; however, the trade-off would be that only a fraction of all patients would receive the additional testing.

The high value of sensitivity found here indicates that the BASH test would have a high rate of “true positives”, i.e., would identify a high proportion (77.8%) of individuals who would screen positive for OSA (according to ODI>=15). The associated 0.646 value of specificity indicates that the BASH test would have a 34.4% rate of “false positives”, i.e., would predict that 34.4% of individuals with low ODI (ODI<15) screen positive for OSA. The “false negative” rate (numerically determined as 1-sensitivity) would be 0.112 i.e., the BASH test would predict that 11.2% of individuals with high ODI (ODI >=15) screen negative for OSA. The desirability and impact of the true positive, false positive, and false negative rates would depend on the proposed process to be followed for individuals with BASH scores above the cutpoint. For example, if the proposed follow-up to BASH>=3 is to determine ODI according to overnight oximetry for a stroke patient, the benefits of detecting possible OSA (i.e., ODI >=15) may outweigh the relatively low risk of measuring overnight oximetry in patients who might screen negative for OSA (i.e., ODI <15). Additional testing and follow-up to confirm presence of OSA in those with ODI>=15 would be needed, to determine the best treatment plan. The proposed BASH tool utilizes simple parameters that are readily available from within electronic medical records (EMRs) and has the potential to be implemented as part of an EMR-embedded clinical decision-making tool for inpatient rehabilitation facilities. The potential gains in post-stroke recovery of function by forestalling the impacts of untreated OSA may warrant such an approach to OSA screening.

## Data Availability

Data may be available per DUA with first author & their institution.

